# Decreased cardio-respiratory information transfer is associated with deterioration and a poor prognosis in critically ill patients with sepsis

**DOI:** 10.1101/2024.08.18.24312167

**Authors:** Cecilia Morandotti, Matthew Wikner, Qijun Li, Emily Ito, Calix Tan, Pin-Yu Chen, Anika Cawthorn, Watjana Lilaonitkul, Ali R Mani

## Abstract

Assessing illness severity in the ICU is crucial for early prediction of deterioration and prognosis. Traditional prognostic scores often treat organ systems separately, overlooking the body’s interconnected nature. Network physiology offers a new approach to understanding these complex interactions. This study used the concept of transfer entropy (TE) to measure information flow between heart rate (HR), respiratory rate (RR), and capillary oxygen saturation (SpO_2_) in critically ill sepsis patients, hypothesizing that TE between these signals would correlate with disease outcome. The retrospective cohort study utilized the MIMIC III Clinical Database, including patients who met Sepsis-3 criteria on admission and had 30 minutes of continuous HR, RR, and SpO_2_ data. TE between the signals was calculated to create physiological network maps. Cox regression assessed the relationship between cardiorespiratory network indices and both deterioration (SOFA score increase of ≥2 points at 48 hours) and 30-day mortality. Among 164 patients, higher information flow from SpO_2_ to HR [TE(SpO_2_→HR)] and reciprocal flow between HR and RR [TE(RR→HR) and TE(HR→RR)] were linked to reduced mortality, independent of age, mechanical ventilation, SOFA score, and comorbidity. Reductions in TE(HR → RR), TE(RR→HR), TE(SpO_2_→RR), and TE(SpO_2_→HR) were associated with increased risk of 48-hour deterioration. After adjustment for potential confounders, only TE(HR→RR) and TE(RR→HR) remained statistically significant. The study confirmed that physiological network mapping using routine signals in sepsis patients could indicate illness severity and that higher TE values were generally associated with improved outcomes.

**New & Noteworthy:** This study adopts an integrative approach through physiological network analysis to investigate sepsis, with the goal of identifying differences in information transfer between physiological signals in sepsis survivors versus non-survivors. We found that greater information flow between heart rate, respiratory rate, and capillary oxygen saturation was associated with reduced mortality, independent of age, disease severity, and comorbidities. Additionally, reduced information transfer was linked to an increased risk of 48-hour deterioration in patients with sepsis.

## Introduction

Sepsis is a complex disease that causes life-threatening organ dysfunction due to a dysregulated host response to infection (Singer et al., 2016). It is one of the most frequent causes of death worldwide, requiring patients to be admitted to intensive care units (ICU) for intensive physiological and clinical monitoring (Rud et al., 2020). The complexity of its pathophysiology and the heterogeneity of its manifestations make sepsis challenging to detect, monitor, and treat. Quantifying illness severity is a crucial aspect of any ICU admission, as it allows for timely interventions to improve outcomes, aids in decision-making, and helps allocate scarce resources (Zimmerman and Kramer, 2014). However, despite the existence of severity scores for almost 40 years, predictions remain imperfect, and they are primarily used for hospital-level case-mix adjustment. Novel digital biomarkers for measuring illness severity may therefore be useful to ICU staff.

The commonest approaches to date have assigned increasing numerical values for progressive dysfunction in each organ system in order to assess their overall association with mortality using regression. Well-known examples include the Sequential Organ Failure Assessment (SOFA) (Vincent et al., 1996), the Simplified Acute Physiology Score II (SAPS II) (Le Gall et al., 1993), the Acute Physiology and Chronic Health Evaluation II (APACHE II) score (Knaus et al., 1985), and the UK’s Intensive Care National Audit and Research Centre (ICNARC) model (Harrison et al., 2007). However, these scores are usually only calculated at the time of critical care admission, or at most on a daily basis, and they often rely on summary measures such as the worst recorded value. Recent machine learning approaches using more granular data have managed to improve short-term prognostication for specific outcomes (Chen et al., 2017; Davies et al., 2020; Henriques et al., 2019; Subramaniam et al., 2014; Yoon et al., 2020), but by treating organs as independent parts to be combined, even these techniques may be ignoring useful information.

Network physiology is a new way of viewing the problem, focussing not on individual organs, but on the degree of interaction *between* them (Bashan et al., 2012). Various measurable aspects of physiology, such as heart rate or respiratory rate, can be conceptualised as “nodes”, with an overall network created by functional connections or “edges” between each node pair if they interact. Strong networks are those which have multiple edges between multiple nodes, or high quantitative values for their connections, as measured by a variety of techniques including simple correlation (Asada et al., 2016, Tan et al., 2020, Zhang et al., 2022) and information transfer (Bartsch et al., 2015; Derakhshan et al., 2019, Jiang et al., 2021). The relevance to illness prediction is that a strong, well-connected network, despite significant individual organ system stress, may represent physiological resilience and predict survival or response to therapy (Asada et al., 2016, Oyelade et al., 2023).

According to information theory, the amount of information in a physiological time series (e.g., heart rate or capillary oxygen saturation fluctuations) can be measured by computing the degree of complexity (i.e., entropy) of the signal (Pincus et al., 1991, Bhogal and Mani, 2017). This idea can be extended to quantify the amount of information exchanged between two physiological signals (Lee et al., 2012, Faes et al.,2014, Jiang et al., 2021). Transfer entropy (TE) is one measure of information transfer between parallel time-series. It is a non-parametric, non-linear extension of the concept of entropy (Schreiber, 2000) that can detect the magnitude and direction of information flow between physiological time series data. TE increases when future values of one time series can be better predicted with knowledge of preceding values from a different time series – suggesting the former is influenced by the latter (Figure 1). One advantage of TE is that it can measure the bidirectional exchange of information between two nodes. For example, it allows us to separately assess how changes in respiratory rate influence capillary oxygen saturation and how changes in oxygen saturation affect respiratory rate. This allows for the assessment of directed interactions between different physiological time series for network mapping based on available physiological signals. For example, using TE, the strength of the cardiorespiratory network could be assessed in experimental hypoxia in healthy participants (Jiang et al., 2021). Assessment of the strength of the cardiorespiratory network in critical illnesses is important in critical care as it may shed light on the pathophysiology of compensatory mechanisms and help predict deterioration or poor outcomes. This is particularly important in complex disorders such as sepsis which is associated with multiorgan failure and high mortality (Srdić et al., 2024, Rud et al., 2020).

**Figure 1.**
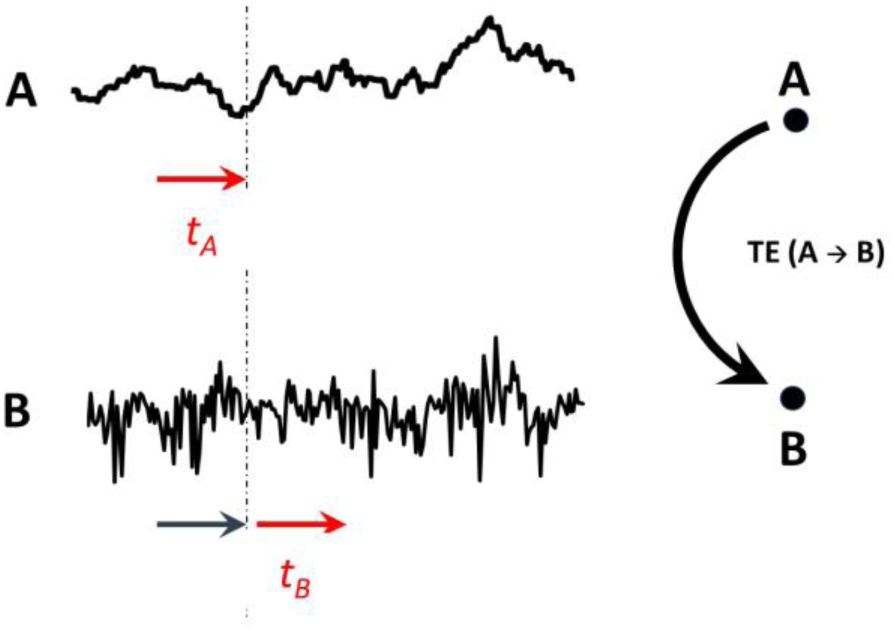
A schematic diagram to explain the concept of transfer entropy (TE). The transfer of information from a physiological time-series A to another parallel time-series B is annotated as TE (A → B) and is defined as how much additional information the past of the A time-series contains about the future observation of the B time-series (red arrows) independently of our knowledge of the past state of B (black arrow). Such transfer of information can be presented as an edge in a network connecting directed information from nodes A to B. *t_A_*: time lag in A from present. *t_B_*: time lag in B from present. As the optimal lag for each node pair is not known a *priori*, TE in this study is measured for a range of time lag values that set equally for both *t_A_* and *t_B_*, at 1, 5, 10, 15, 20 and 25 seconds.

This study therefore aimed to understand whether cardiorespiratory transfer entropy, measured from bedside monitor data of patients with sepsis in the MIMIC-III database, could be used to assess their physiological network strength and its relationship with 48-hour deterioration and mortality.

## Materials and methods

This was a retrospective cohort study using the Waveform Database Matched Subset of the Medical Information Mart for Intensive Care III (MIMIC-III) Clinical Database (Johnson et al., 2016; Moody et al., 2017), reported in accordance with the RECORD guidelines (Benchimol et al., 2015).

### Ethics statement

The MIMIC-III was anonymized following HIPAA standards and the project received approval from the Institutional Review Boards of Beth Israel Deaconess Medical Center and MIT (IRB protocol nos. 2001P001699/14 and 0403000206, respectively; Johnson et al., 2016. The authors who handled the data underwent required ethics training at MIT and were credentialed (ID 10304625).

### Participants and data extraction

Details of patient enrolment flow diagram and data extraction is described elsewhere (Gheorghita et al., 2022). In brief, inclusion was limited to patients over 18 years of age, with a single ICU stay who met the Sepsis-3 criteria on admission (an increase in SOFA score of >= 2 points and suspicion of infection (Singer et al., 2016)). To ensure adequate data for stable estimation of TE, patients were only included if their waveform record contained at least 30 minutes continuous and simultaneous time series data when resampled to 1Hz (Ito et al., 2011; Jiang et al., 2021). 179 records met these criteria when considering three waveforms - heart rate (HR), respiratory rate (RR) and capillary oxygen saturation (SpO_2_) – and formed the basis of the final cohort. Matched information was retrieved from the Clinical Database on patient age, sex, SOFA scores, Elixhauser comorbidity index, mechanical ventilation, and date of death. A 30-day survival data was missing in 15 patients; therefore, 164 patients were included in the final survival analysis

### Definition of deterioration

The SOFA score was extracted for the day when a patient’s physiological signals record was available and again 48 hours later. Deterioration was defined as SOFA score >= 2 points at 48 hours. Due to early discharge or death, 55% of 48-hour SOFA scores in this study required imputation. To handle missing for data for 48 hour SOFA scores calculation, the maximum score of 24 was applied if the patient had already died (Lambden et al., 2019), and a score of 1 applied if discharged alive from ICU.

### Calculation of transfer entropy (TE)

An existing open-source algorithm (https://www.physionet.org/content/tewp/1.0.0/) was used to calculate TE (in bits) for parallel physiological time-series. This algorithm employs an extension of Darbellay-Vajda adaptive partitioning (Lee et al., 2012) to estimate a non-linear probability density function in a computationally efficient manner. It calculates the probability of event B of a time lag window length of t_B_ occurring after the outcome of event A of a time lag of window length of t_A_ was observed (Figure 1), where A and B are representations of the physiological parameters (e.g., HR, RR, and SpO_2_). The returned value of transfer entropy represents the amount of directional information transferred from a data segment of one physiological time series to the future data segment of another time series. In addition to probability density function estimation, TE magnitude also depends on the lag chosen between the source and target time series. As the optimal lag for each node pair was not known *a priori*, TE was measured for a range of time lag values that set equally for both t_A_ and t_B_, at 1, 5, 10, 15, 20 and 25 seconds (Figure 1). This approach was conducted to ascertain the consistency of the results and establish an optimal time lag for future transfer entropy computations. Based on these results, a time lag value of 5 seconds was chosen to calculate the TE estimate for each edge for all patients.

### Network visualization

Network maps were constructed for qualitative assessment by conceptualising each physiological signal as a node, with edges drawn between nodes showing the strength of any directional information flow. TE edge strength was displayed as the average group value. Comparison of directed transfer entropy values between physiological time-series were conducted at a time lag of 5 seconds, with each mean transfer entropy calculation being compiled to form an adjacency matrix. This matrix was then used to plot a bidirectional network graph in MATLAB.

### Centrality measure

Centrality is a measure of the importance of a node within a network in terms of flow of information. Indegree (ID) and outdegree (OD) measure the centrality of a node by calculating the information that each node receives (ID) or sends out (OD). Indegree and outdegree centralities of SpO_2_, HR, and RR were calculated for each patient using respective transfer entropy adjacency matrices using MATLAB.

### Statistical analysis

Data are shown as mean ± SD unless stated otherwise. The mean differences in network edges and node centralities between the groups (survivors vs non-survivors and deterioration vs no deterioration) were calculated using the Student’s t-test or its non-parametric equivalent (Mann-Whitney U-test).

Cox regression was used for estimation of hazard ratios with 95% confidence intervals. Multivariate Cox regression was performed with covariates of SOFA, mechanical ventilation, Elixhauser comorbidity score, and age. ROC curve analysis was used to find optimum cut-off point (Youden’s index) with optimum sensitivity and specificity in prediction of 30-day mortality in the intensive care unit and of deterioration. To visualise patient survival, the Kaplan Meier curves were applied and analysed using a log rank (Mantel-Cox) method. P-value less than 0.05 was used for statistical significance. Two-way ANOVA was used for assessment of the effect of time lag on TEs. We also wondered if shorter time-series (namely, 20, 10, 5, 2 and 1-min) can estimate TE calculated from 30-min time-series and predicts poor outcomes (mortality, deterioration) within this patient population. Thus, Bland-Altman plots were used to identify bias in TE of time series of 20, 10, 5, 2, and 1 minutes (starting from the beginning of recording) compared to the 30-min transfer entropy values. This method is based on the quantification of the agreement between two quantitative measurements (short time-series, A versus 30-min, B) by studying the relationship between *A* − *B* and (*A* + *B*)/2 (Bland and Altman, 1999). The linear regression analysis was used to test for statistical significance of the bias for the intercept and slope in the Bland-Altman plots (Giavarina, 2015).

## Results

Descriptive characteristics of the participants are shown in Table 1. Overall, 130 patients survived after a 30-day follow-up period. The non-survivors (n = 34) were older (65 ± 18 vs.75 ± 12, P=0.003) and had higher SOFA scores (4.1 ± 2.3 vs. 6.8 ± 4.1, P<0.001). The comorbidity index (Elixhauser) was higher in non-survivors (P=0.027). Changes in SpO_2_ mean and pattern of fluctuations in this cohort has been reported elsewhere (Gheorghita et al., 2022). In brief, the average SpO_2_ was marginally higher in the survivors compared to the non-survivors (97.4 ±2.2 vs. 96.0 ± 6.3, P = 0.033). Mean HR was higher in survivors compared to the non-survivors (83.5 ± 18.3 vs. 94.0 ± 23.8 beats/min, P = 0.0063)). There was no statistical difference in RR between survivors and non-survivors (19.7 ± 4.8 vs. 21.2 ± 6.0 breath/min, P=0.117). There was no difference in distribution of gender of ethnicity between survivors and non-survivors.

**Table 1.** Summary of descriptive results.

| Variable | Summary value |
| --- | --- |
| Age (years) (median, Interquartile range) | 68 (53-84) |
| Male/Female (count, %) | 93/71 (57%/43%) |
| Ethnicity (count, %) |  |
| White | 125 (76%) |
| Black | 10 (6%) |
| Hispanic | 5 (3%) |
| Asian | 3 (1%) |
| Other | 3 (1%) |
| Unknown | 18 (11%) |
| Hospital admission primary diagnosis (count, %) |  |
| Neurological | 46 (28%) |
| Cardiac | 35 (21%) |
| Infective | 34 (21%) |
| Gastrointestinal | 9 (5%) |
| Orthopaedic | 4 (2%) |
| Other | 35 (21%) |
| Elixhauser index (median, Interquartile range) | 3 (0-7) |
| Mechanically ventilated during TE measurement (count, %) | 45 (27.4%) |
| SOFA score on day of TE was measured (median, Interquartile range) | 4 (2-6) |
| SOFA score 48 hours after TE was measured (median, Interquartile range) | 1 (1-5) |
| Deterioration in SOFA score $\geq 2$ points at 48 hours (count, %) | 31 (18.9%) |
| 30-day mortality (count, %) | 34 (20.7%) |

Association of transfer entropy and network indices with 30-day mortality The TE values are subsequently denoted as follows:

|  |  |
| --- | --- |
| From heart rate to respiratory rate: | TE (HR → RR) |
| From heart rate to oxygen saturation: | TE (HR → SpO<sub>2</sub>) |
| From respiratory rate to heart rate: | TE (RR → HR) |
| From respiratory rate to oxygen saturation: | TE (RR → SpO<sub>2</sub>) |
| From oxygen saturation to heart rate: | TE (SpO<sub>2</sub> → HR) |
| From oxygen saturation to respiratory rate: | TE (SpO<sub>2</sub> → RR) |

As shown in Table 2, the highest average value of TE was during the transfer of information from SpO_2_ to RR. The lowest TE between physiological signals was during the transfer of information from HR to SpO_2_. TE values in most directions were significantly higher in survivors compared to non-survivors after 30 days of follow-up (Table 2).

**Table 2:**
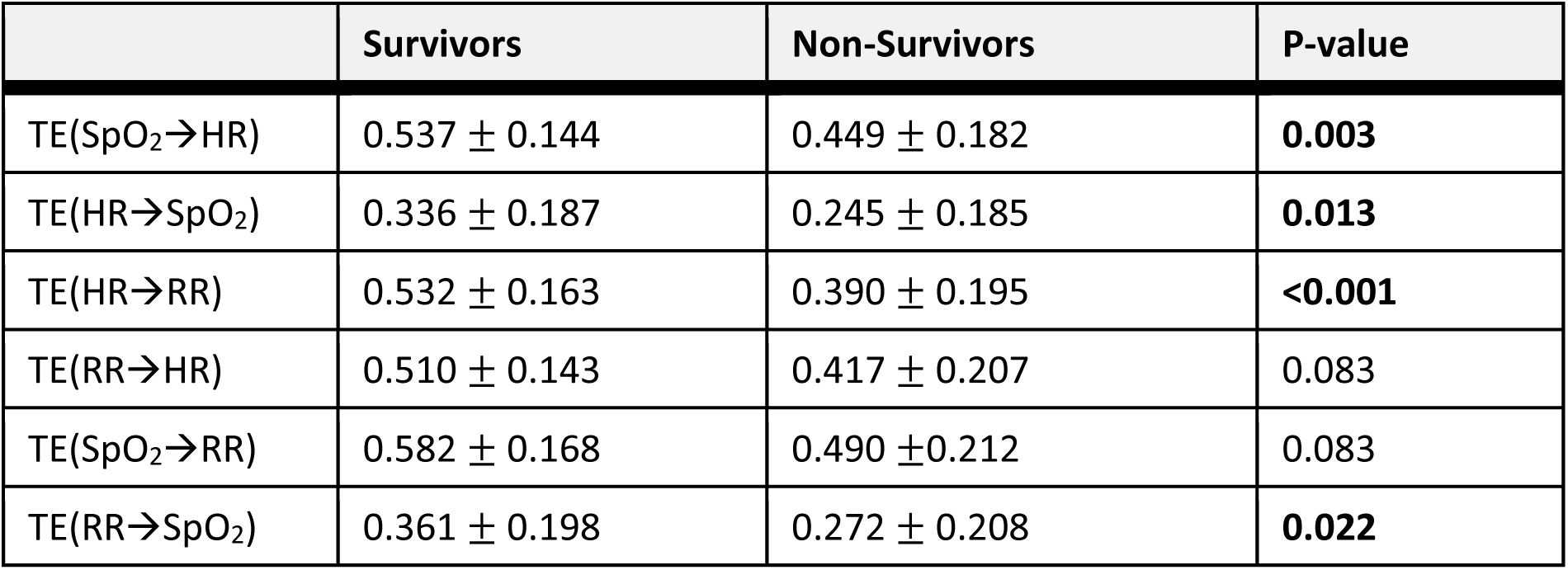
Comparison of Transfer Entropy Means between Survivors and Non-survivors:

|  | Survivors | Non-Survivors | P-value |
| --- | --- | --- | --- |
| TE(SpO<sub>2</sub>→HR) | 0.537 ± 0.144 | 0.449 ± 0.182 | **0.003** |
| TE(HR→SpO<sub>2</sub>) | 0.336 ± 0.187 | 0.245 ± 0.185 | **0.013** |
| TE(HR→RR) | 0.532 ± 0.163 | 0.390 ± 0.195 | **<0.001** |
| TE(RR→HR) | 0.510 ± 0.143 | 0.417 ± 0.207 | 0.083 |
| TE(SpO<sub>2</sub>→RR) | 0.582 ± 0.168 | 0.490 ± 0.212 | 0.083 |
| TE(RR→SpO<sub>2</sub>) | 0.361 ± 0.198 | 0.272 ± 0.208 | **0.022** |

### Centrality measures

To assess the importance of each node within the network, centrality indexes (indegree and outdegree) were measured and compared between the groups. The results indicate that within the HR-RR-SpO_2_ network, the RR node receives the highest amount of information from other nodes (highest indegree), and the SpO_2_ node sends the highest amount of information to other nodes (highest outdegree). Table 3 shows details of the centrality measures between groups. There is a significant difference between survivors and non-survivors in centrality measures indicating that all nodes have higher centrality in survivors compared with non-survivors.

**Table 3:**
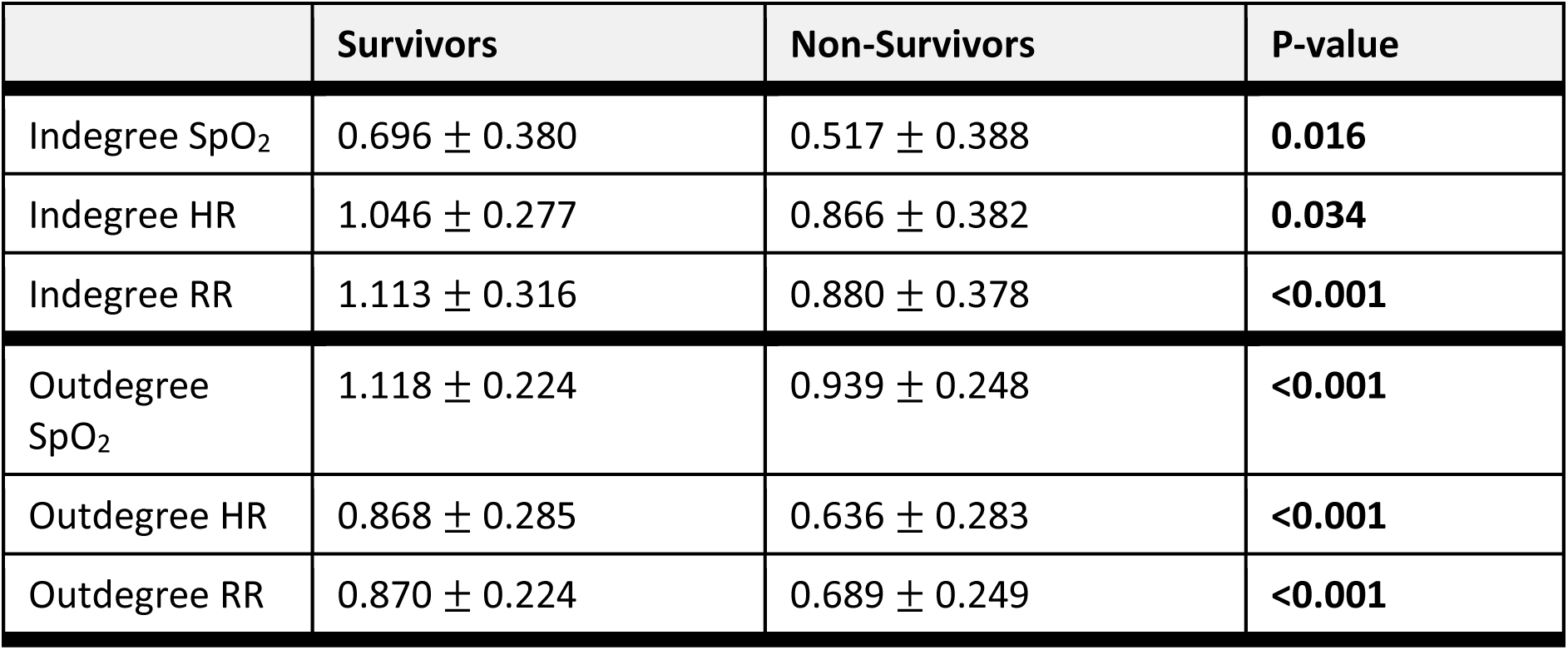
Comparison of Centrality Indices (Indegree and Outdegree) between Survivors and Non-Survivors.

|  | Survivors | Non-Survivors | P-value |
| --- | --- | --- | --- |
| Indegree SpO<sub>2</sub> | 0.696 ± 0.380 | 0.517 ± 0.388 | **0.016** |
| Indegree HR | 1.046 ± 0.277 | 0.866 ± 0.382 | **0.034** |
| Indegree RR | 1.113 ± 0.316 | 0.880 ± 0.378 | **<0.001** |
| Outdegree SpO<sub>2</sub> | 1.118 ± 0.224 | 0.939 ± 0.248 | **<0.001** |
| Outdegree HR | 0.868 ± 0.285 | 0.636 ± 0.283 | **<0.001** |
| Outdegree RR | 0.870 ± 0.224 | 0.689 ± 0.249 | **<0.001** |

### Survival analysis

Cox regression analysis was conducted to evaluate the risk of 30-day mortality associated with TE and centrality measures (Table 4). Reduction in TE or centrality of individual nodes were associated with increased chance of mortality in this cohort of patients with sepsis. Since non-survivors were older and had higher SOFA scores and comorbidities, we considered whether these characteristics might confound the association between TE and mortality. Additionally, factors such as mechanical ventilation could affect physiological time-series data and potentially influence these findings. To address these concerns, we performed multivariate Cox regression analysis to assess the dependence of individual network indices on age, SOFA score, comorbidity index (Elixhauser), and mechanical ventilation. The results indicated that among network indices, TE (SpO_2_ → HR), TE (HR → RR), TE (RR → HR), Indegree of HR and all outdegrees (HR, RR and SpO_2_) were independent predictors of 30-day mortality (Supplementary material 1). Lower TE values in the group that went on to die, suggests reduced connectivity and weakened cardiorespiratory network in non-survivor. Graphical visualization of these network edges is shown in Figure 2.

**Figure 2.**
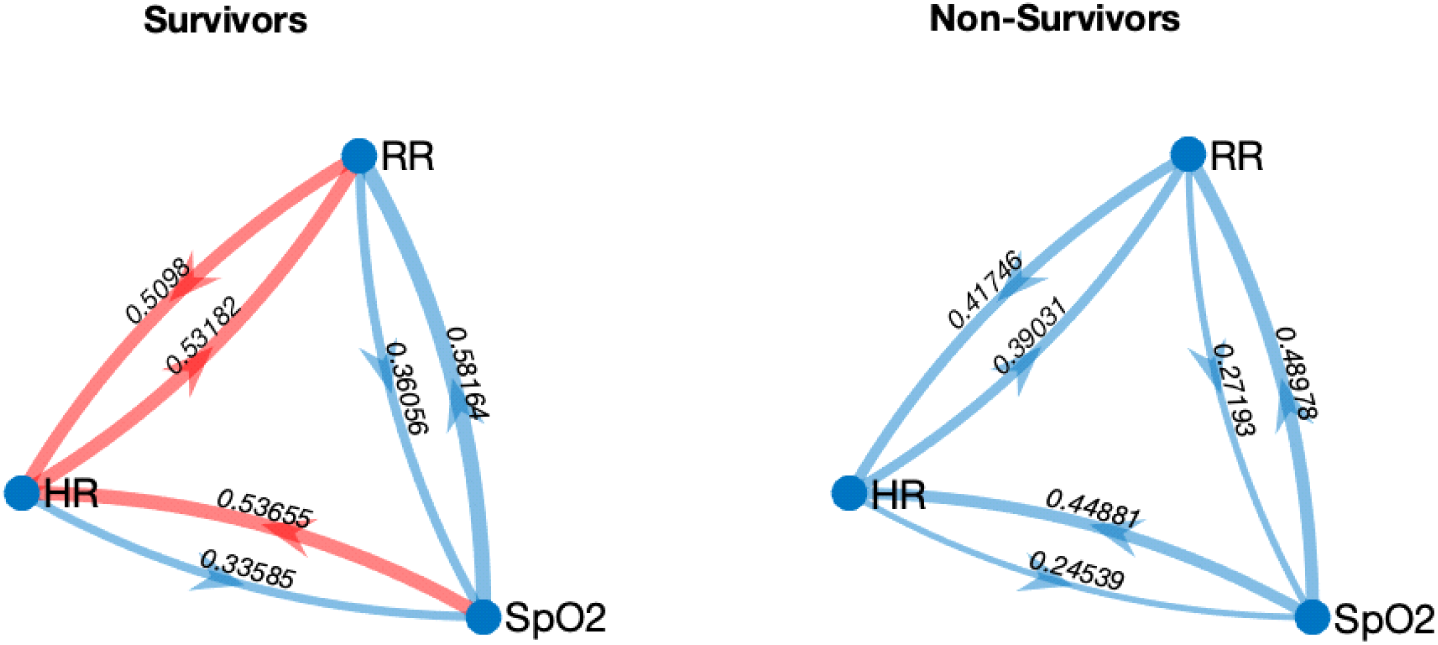
Network maps for survivors and non-survivors, showing mean TE values (in bits) for each edge. Red: TEs which are significant predictors of mortality, independent of covariates (age, SOFA, comorbidity index and mechanical ventilation). Edge weighting correspond to magnitude of information flow. HR: heart rate; RR: respiratory rate; SpO_2_: oxygen saturation.

**Table 4:** Monovariate Cox regression analysis to predict 30-day mortality based on Transfer Entropies, and Network Centrality Indices (Indegrees and Outdegrees):

|  | B | SE | P-value | Exp(B) | Confidence Interval (95%) |
| --- | --- | --- | --- | --- | --- |
| TE(SpO<sub>2</sub>→HR) | -2.633 | 0.901 | **0.003** | 0.072 | 0.012 – 0.421 |
| TE(HR→SpO<sub>2</sub>) | -2.070 | 0.881 | **0.019** | 0.126 | 0.022 – 0.709 |
| TE(HR→RR) | -3.357 | 0.789 | **<0.001** | 0.035 | 0.007 – 0.166 |
| TE(RR→HR) | -2.694 | 0.898 | **0.003** | 0.068 | 0.012– 0.393 |
| TE(SpO<sub>2</sub>→RR) | -2.122 | 0.763 | **0.005** | 0.120 | 0.027 – 0.535 |
| TE(RR→SpO<sub>2</sub>) | -1.765 | 0.821 | **0.032** | 0.171 | 0.034 – 0.856 |
| Indegree SpO<sub>2</sub> | -0.965 | 0.426 | **0.023** | 0.381 | 0.165 – 0.877 |
| Indegree HR | -1.382 | 0.453 | **0.002** | 0.251 | 0.103 – 0.610 |
| Indegree RR | -1.421 | 0.388 | **<0.001** | 0.242 | 0.113 - 0.517 |
| Outdegree SpO<sub>2</sub> | -2.476 | 0.629 | **<0.001** | 0.084 | 0.025 – 0.289 |
| Outdegree HR | -2.015 | 0.511 | **<0.001** | 0.133 | 0.049 – 0.363 |
| Outdegree RR | -2.629 | 0.693 | **<0.001** | 0.072 | 0.019 – 0.281 |

### The effect of time lag on transfer entropy

To ensure that an optimized time lag value is used for TE calculation, TE was measured for a range of time lag values at 1, 5, 10, 15, 20 and 25 second. Survivor group consistently had higher transfer entropy values at all time lags (Figure 3). It is noteworthy that when the calculation was set between a time lag of 5 and 25, the resulting transfer entropy values fell within a comparable range, as opposed to when a time lag of 1 was utilized. This substantiates the use of time lag 5 seconds in transfer entropy calculation. TE (SpO_2_ → HR), TE (HR → RR) and TE (RR → HR) were chosen for this analysis as they demonstrated a significant predictive power in multivariate Cox regression analysis for mortality.

**Figure 3:**
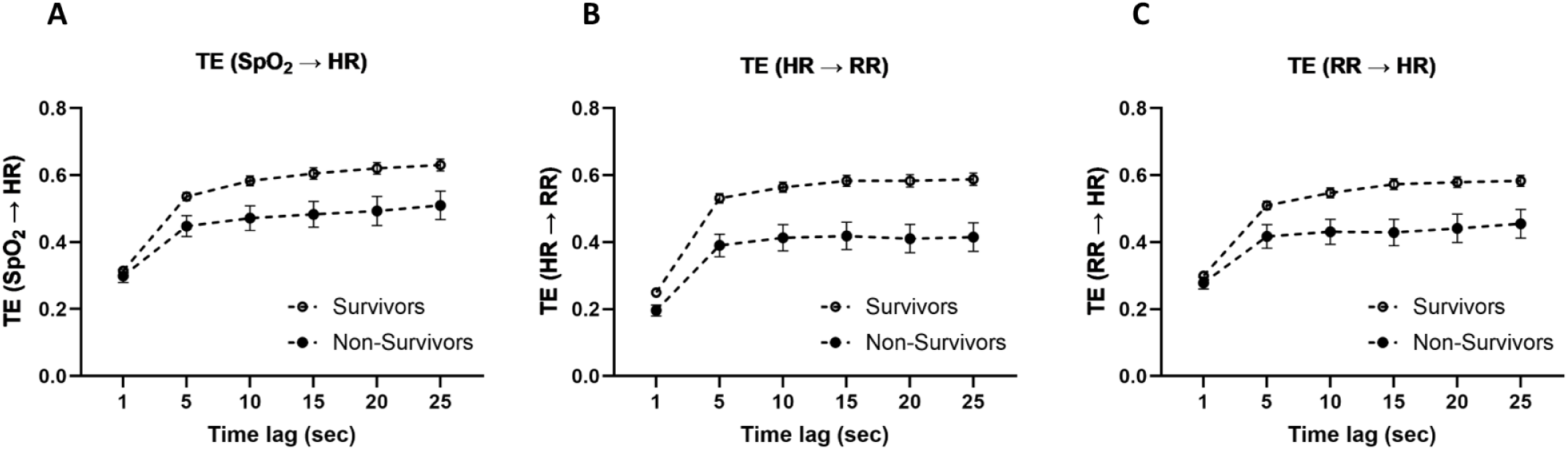
Comparison of Transfer Entropies [TE (SpO_2_ → HR), TE (HR → RR) and TE (RR → HR)] between survivors and non-survivors at different time lags. Data are shown as mean standard error or mean. Two-way ANOVA showed that group (Survivors/Non-Survivors) and time lag both significantly affect TEs (P<0.001 for all TEs) and there is no interaction between group and time-lag.

### Association of transfer entropy and network indices with 48-hours deterioration

31 (18.9%) patients had an increase in SOFA score >= 2 points at 48 hours. TE values and network indices of this group were compared with the rest of the patients who didn’t show 48-hour deterioration. As shown in Table 5, TE (SpO_2_ → HR), TE (HR → RR) and TE (RR → HR) were significantly lower in the group that exhibited deterioration. Likewise, the centrality measures of all nodes, except for indegree SpO_2_, were significantly lower in the deteriorating group.

**Table 5:** Comparison of Transfer Entropy Means between Patients with 48-hour Deterioration and without Deterioration.

|  | Deterioration | No Deterioration | P-value |
| --- | --- | --- | --- |
| TE($\text{SpO}_2 \rightarrow \text{HR}$) | $0.451 \pm 0.173$ | $0.534 \pm 0.149$ | **0.007** |
| TE($\text{HR} \rightarrow \text{SpO}_2$) | $0.282 \pm 0.180$ | $0.325 \pm 0.191$ | 0.247 |
| TE($\text{HR} \rightarrow \text{RR}$) | $0.391 \pm 0.211$ | $0.528 \pm 0.161$ | **0.005** |
| TE($\text{RR} \rightarrow \text{HR}$) | $0.428 \pm 0.195$ | $0.505 \pm 0.151$ | **0.016** |
| TE($\text{SpO}_2 \rightarrow \text{RR}$) | $0.479 \pm 0.234$ | $0.582 \pm 0.161$ | 0.231 |
| TE($\text{RR} \rightarrow \text{SpO}_2$) | $0.325 \pm 0.213$ | $0.346 \pm 0.200$ | 0.590 |

### Survival analysis

Cox regression analysis showed that reduction in most TE or centrality of individual nodes were associated with increased chance of 48-hour deterioration in this cohort of patients with sepsis (Table 7). However, after controlling for age, SOFA, Elixhauser comorbidity index and mechanical ventilation, only TE (HR → RR) and TE (RR → HR) remained statistically significant suggesting that these edges provide information on 48-hour deterioration independent of other clinical covariates. A summary of multivariate Cox regression analysis is shown in Supplementary material 2. Graphical visualization of these network edges is shown in Figure 4.

**Figure 4.**
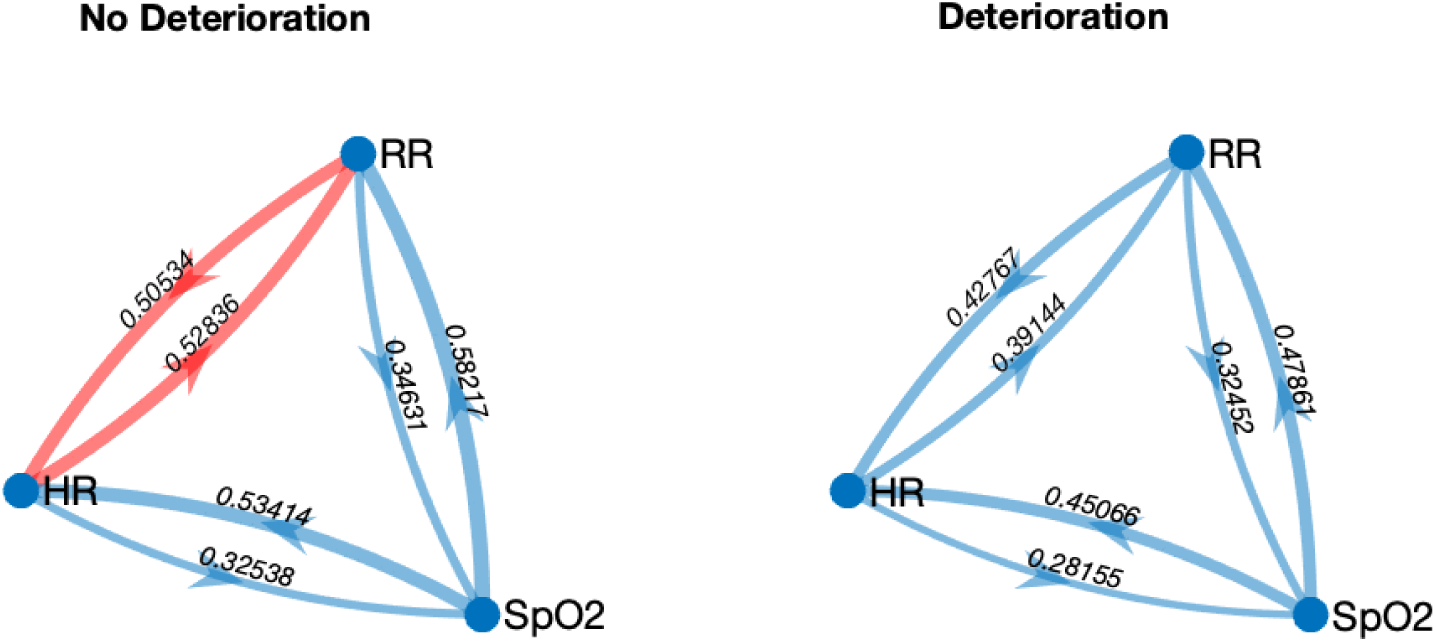
Network maps for patients with 48-hour deterioration and no deterioration, showing mean TE values (in bits) for each edge. Edge weighting correspond to magnitude of information flow. HR: heart rate; RR: respiratory rate; SpO_2_: oxygen saturation.

**Table 6:** Comparison of Centrality Indices (Indegree and Outdegree) between Patients with 48-hour Deterioration and without Deterioration.

|  | **Deterioration** | **No Deterioration** | **P-value** |
| --- | --- | --- | --- |
| Indegree SpO<sub>2</sub> | 0.606 ± 0.388 | 0.672 ± 0.388 | 0.398 |
| Indegree HR | 0.878 ± 0.365 | 1.0395 ± 0.288 | **0.009** |
| Indegree RR | 0.870 ± 0.426 | 1.11 ± 0.304 | **0.017** |
| Outdegree SpO<sub>2</sub> | 0.929 ± 0.268 | 1.116 ± 0.220 | **<0.001** |
| Outdegree HR | 0.673 ± 0.312 | 0.854 ± 0.286 | **0.002** |
| Outdegree RR | 0.752 ± 0.241 | 0.852 ± 0.237 | **0.038** |

**Table 7:**
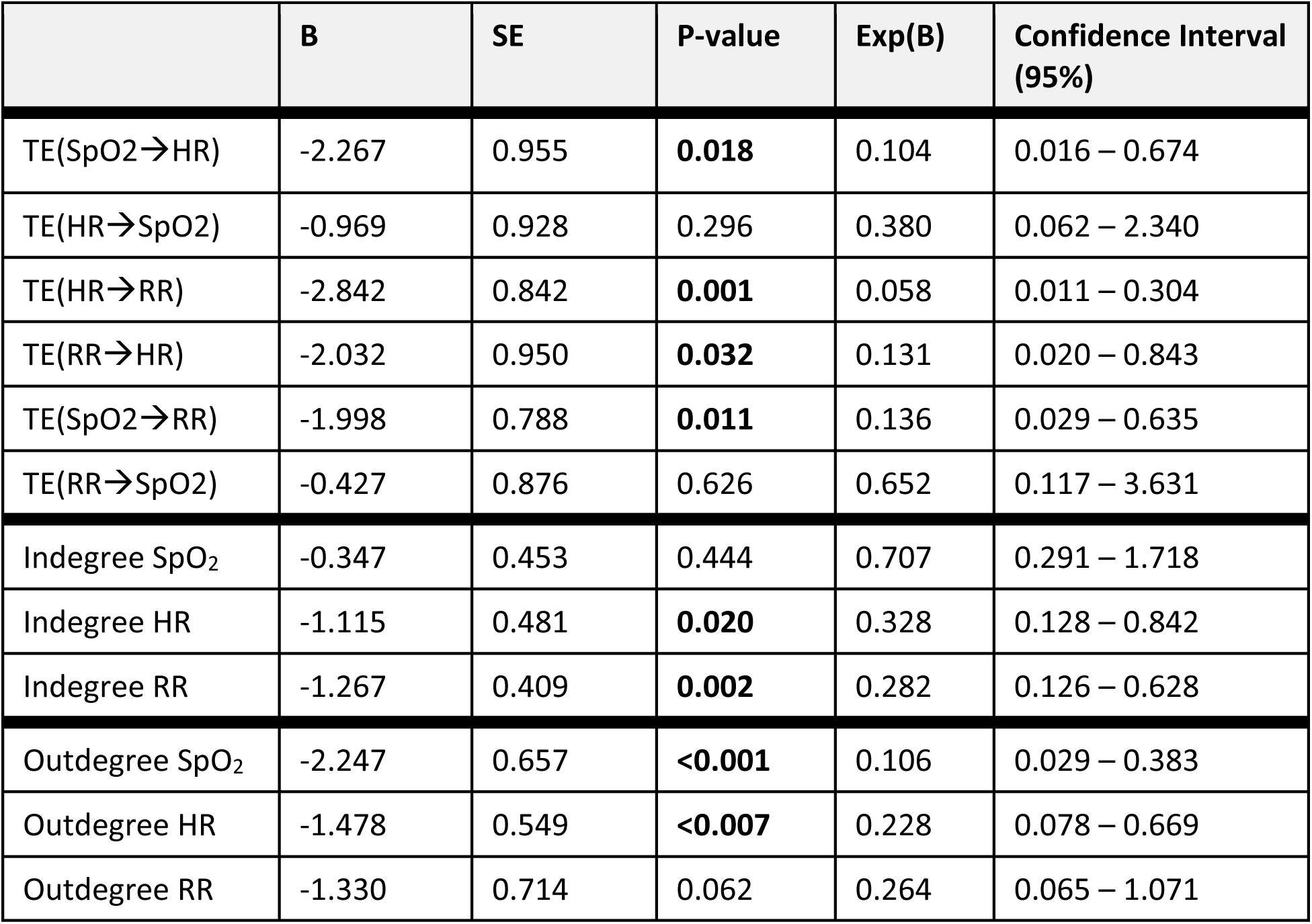
Monovariate Cox regression analysis to predict 48-hour Deterioration based on Transfer Entropies, and Network Centrality Indices (Indegrees and Outdegrees)

|  | **B** | **SE** | **P-value** | **Exp(B)** | **Confidence Interval (95%)** |
| --- | --- | --- | --- | --- | --- |
| TE(SpO<sub>2</sub>→HR) | -2.267 | 0.955 | **0.018** | 0.104 | 0.016 – 0.674 |
| TE(HR→SpO<sub>2</sub>) | -0.969 | 0.928 | 0.296 | 0.380 | 0.062 – 2.340 |
| TE(HR→RR) | -2.842 | 0.842 | **0.001** | 0.058 | 0.011 – 0.304 |
| TE(RR→HR) | -2.032 | 0.950 | **0.032** | 0.131 | 0.020 – 0.843 |
| TE(SpO<sub>2</sub>→RR) | -1.998 | 0.788 | **0.011** | 0.136 | 0.029 – 0.635 |
| TE(RR→SpO<sub>2</sub>) | -0.427 | 0.876 | 0.626 | 0.652 | 0.117 – 3.631 |
| Indegree SpO<sub>2</sub> | -0.347 | 0.453 | 0.444 | 0.707 | 0.291 – 1.718 |
| Indegree HR | -1.115 | 0.481 | **0.020** | 0.328 | 0.128 – 0.842 |
| Indegree RR | -1.267 | 0.409 | **0.002** | 0.282 | 0.126 – 0.628 |
| Outdegree SpO<sub>2</sub> | -2.247 | 0.657 | **<0.001** | 0.106 | 0.029 – 0.383 |
| Outdegree HR | -1.478 | 0.549 | **<0.007** | 0.228 | 0.078 – 0.669 |
| Outdegree RR | -1.330 | 0.714 | 0.062 | 0.264 | 0.065 – 1.071 |

### Diagnostic performance of network indices for 30-day mortality

ROC curve analysis was performed to evaluate diagnostic performance of TEs and centrality measures for mortality (Figure 5), where TE (HR → RR) and outdegree HR showed the highest accuracy for sensitivity and specificity than the other classifiers (AUC > 0.5, P<0.01 for all variables).

**Figure 5.**
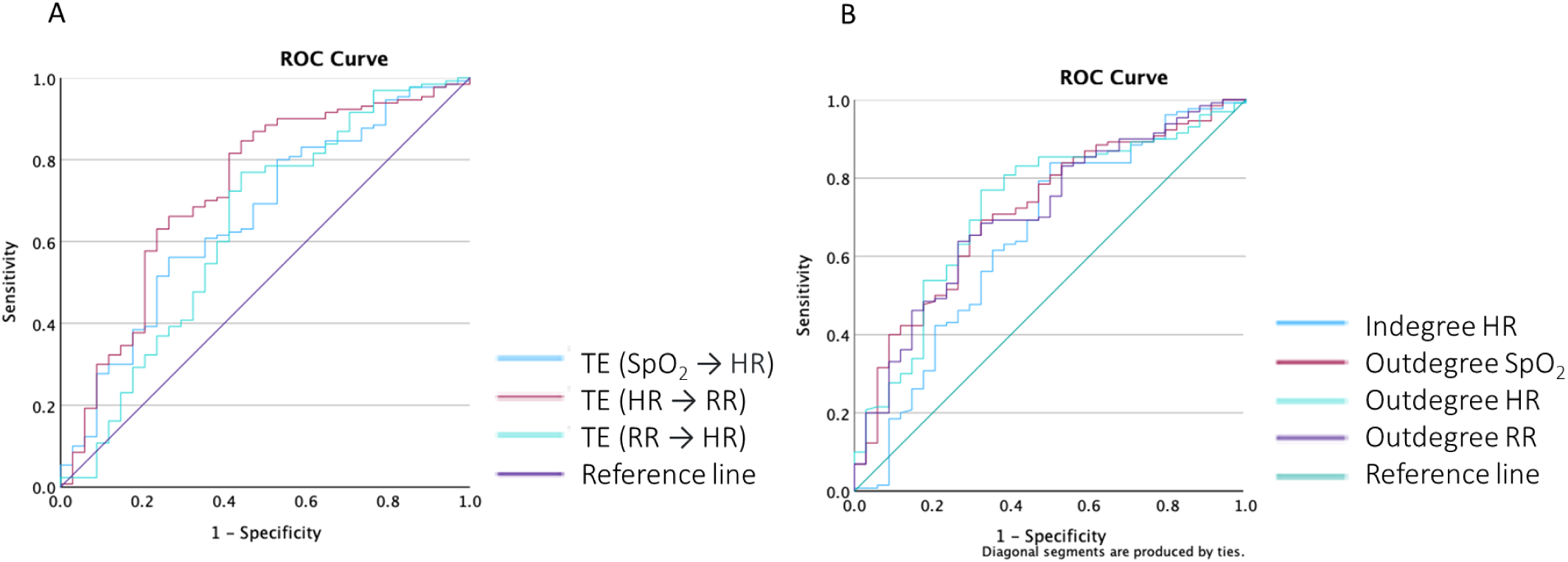
ROC Curves for Prediction of 30-day mortality based on Transfer Entropies (A) and Centrality Indices (B)

Kaplan-Meier survival plots were constructed to compare survival between different directional TE groups and between outdegrees of TE, categorised based on the Youden index threshold of ROC for 30-day mortality. Kaplan-Meier plots (Figure 6 and 7) showed separation of these groups’ survival curves based on the thresholds for TE (SpO_2_ → HR), TE (HR → RR), TE (RR → HR) and all outdegrees with statistical significance assessed using the log rank test (p < 0.001). Indegree of HR is also a significant predictor of mortality in the log rank test. Data not shown.

**Figure 6:**
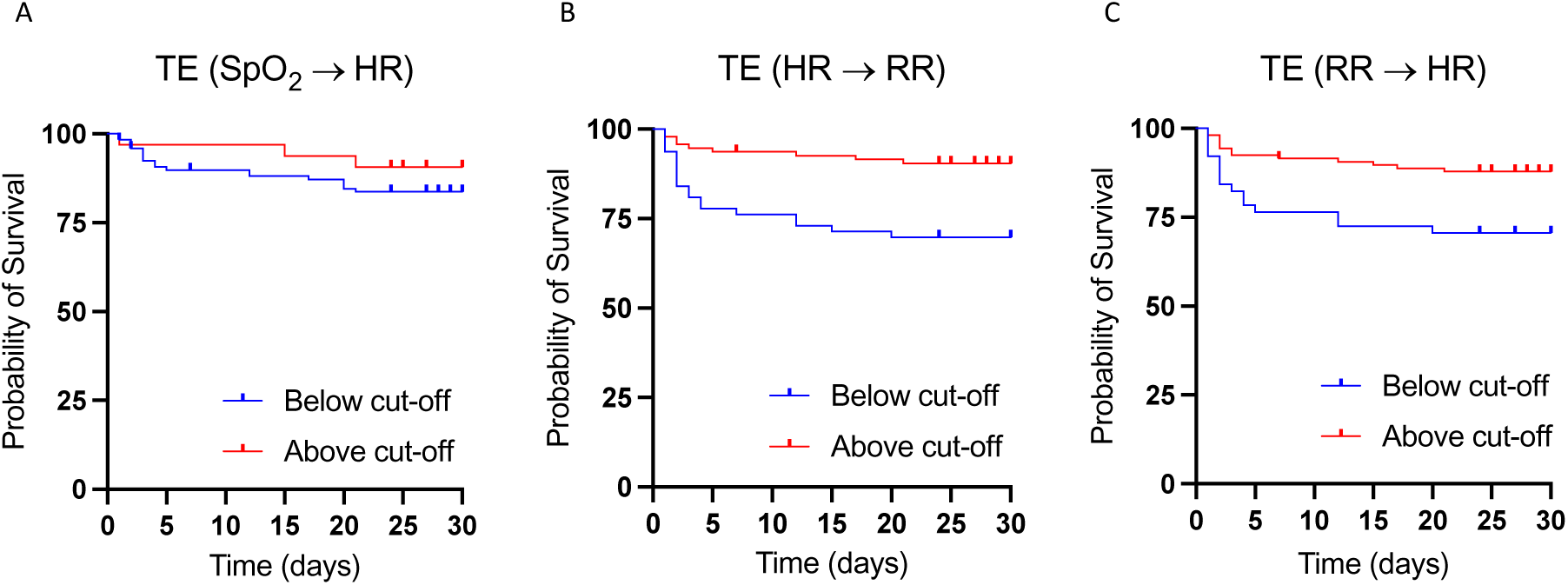
Kaplan Meier Graphs for Visualization of Prediction of Mortality based on Transfer Entropies. ROC curves were used to obtain optimum cut-off points.

**Figure 7:**
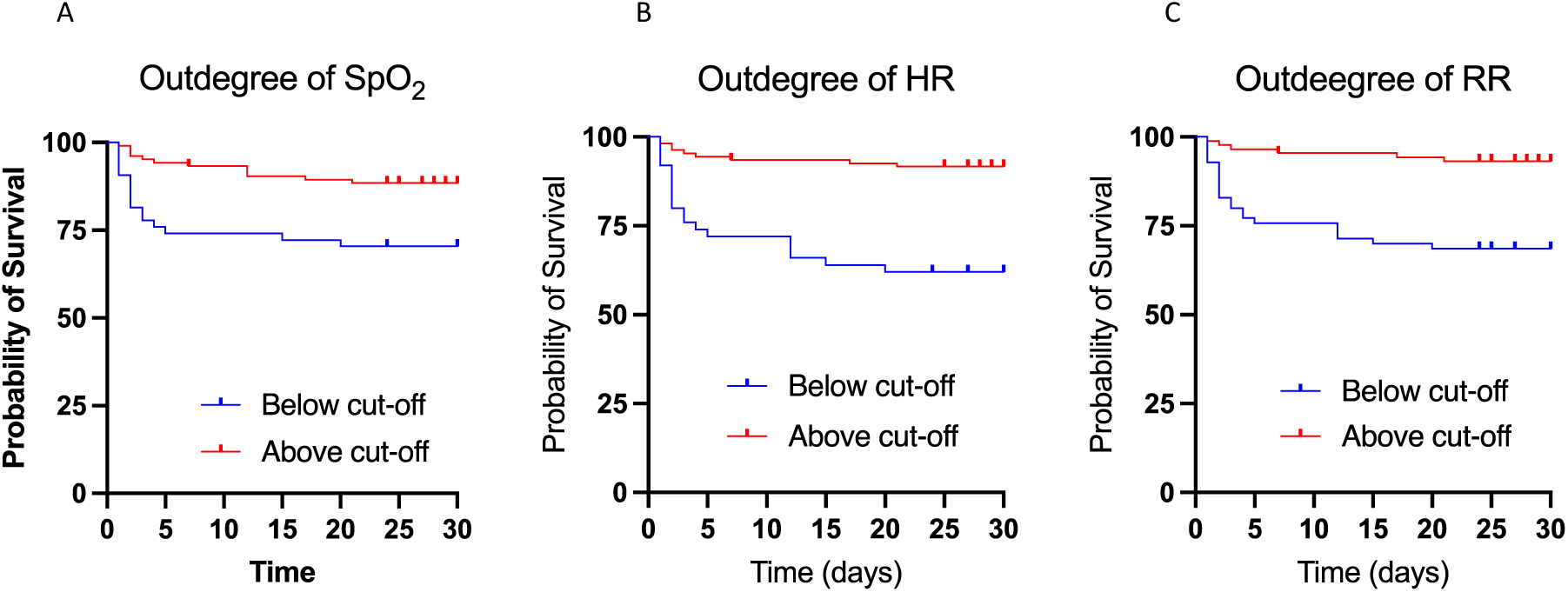
Kaplan Meier Graphs for Visualization of Prediction of Mortality based on Centrality Indices. ROC curves were used to obtain optimum cut-off points.

## Discussion

This study takes an integrative approach through network analysis to investigate sepsis and aims to identify differences in the information transfer and connectivity of organ systems between sepsis survivors and non-survivors. To optimize the mapping method’s network analysis, we investigated the suitable range of time lag for transfer entropy calculation. Transfer entropy has rarely been applied in sepsis prognosis or organ deterioration assessment, even though HR, RR, and SpO_2_ signals are closely monitored in clinical settings, and transfer entropy calculation has a well-established algorithm. Using HR, RR, and SpO_2_ as clinical variables to represent the cardio-respiratory system, the study investigated the transfer entropy values of 164 sepsis patients in the ICU.

### Summary of results and interpretation

This study demonstrated several important new findings:

Firstly, the study found that the group means of all transfer entropy values were significantly higher in survivors than in non-survivors, indicating more active physiological systems and greater information transfer in patients with better prognoses. This supports the hypothesis that decreased homeostatic interorgan connectivity is associated with poor prognosis in critically ill sepsis patients, which is also consistent with previous studies on organ systemic dysfunction in critically ill patients (Asada et al., 2016) and patients with cirrhosis (Tan et al., 2020). In normal health, heart rate, cardiac output, blood pressure, respiratory rate, tidal volume and many other measurable aspects of cardiorespiratory physiology are intricately linked via positive and negative feedback systems. Exactly how mutual effects are mediated is still not perfectly understood (Guyenet, 2014), but increases in blood pressure stimulate arterial baroreceptors, leading to slowing of respiration (West and Luks, 2020), and changes in arterial oxygen saturation can similarly be precipitated by changes in the cardiovascular system, as these affect arterial oxygen tension via altered ventilation-perfusion matching in the lung. The mechanism and benefits of respiratory sinus arrhythmia (changes in HR in each respiratory cycle) is well documented (Ben-Tal et al., 2012). The effect of RR on HR and blood pressure, via changes in intrathoracic pressure, is already used widely in anaesthesia and intensive care medicine to understand intravascular volume status (Vistisen et al., 2019). There is also a wealth of evidence showing that heart rate variability (HRV) is lower in patients with worse ICU outcomes (Karmali et al., 2017), something which would be consistent with partial uncoupling of organ-systems and reduced TE in pairs that included heart rate. Likewise, reduced oxygen saturation entropy has recently been reported in non-surviving patients with sepsis (Gheorghita et al, 2022) which is line with reduced transfer of information between nodes that included SpO_2_ in patients with poor prognosis. Reduced transfer of information between physiological signals may represent uncoupling of organ systems during a pathologic challenge (e.g. infection). While it is expected that compensatory mechanisms lead to enhanced coupling of physiological subsystems during physiologic challenges, in the group of patients who have uncoupled physiological networks, this may lead to deterioration and death (Figure 8). The reason behind the uncoupling of organ systems in life-threatening sepsis is not well understood. Experimental reports suggest end-organ hypo-responsiveness to autonomic neural stimulation (Hajiasgharzadeh et al., 2011; Gholami et al., 2012), decreased controllability of the cardiac pacemaker (Mazloom et al., 2014), and/or impaired neural processing within the brainstem autonomic regulatory centres (e.g., the Nucleus of the Solitary Tract) (Eftekhari et al., 2020) during experimental sepsis.

**Figure 8.**
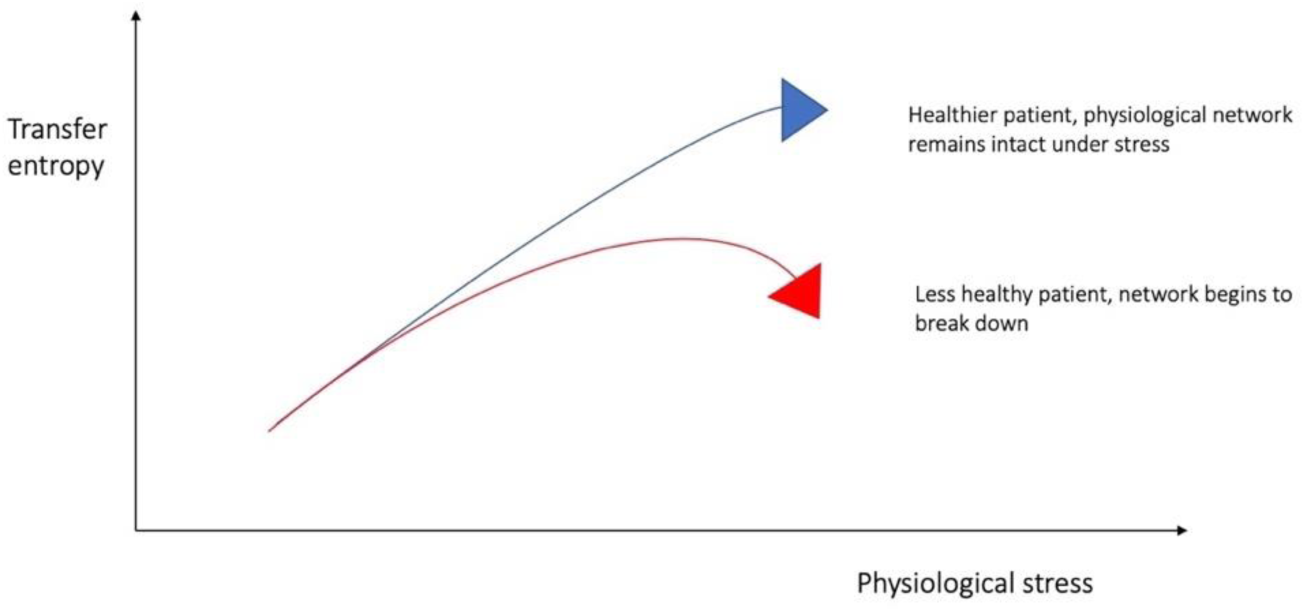
Graphical representation of possible underlying relationship between physiological stress and TE

Secondly, the study demonstrated that directed transfer entropy from physiological time-series can predict mortality and 48-hour organ function deterioration in critically ill patients with sepsis, independent of SOFA score, comorbidity and ventilation status. These findings highlight the potential of transfer entropy in filling the gap in foreseeing the potential underlying dysfunctional connections between organ systems of complex diseases. Measurement of HR, RR, and SpO_2_ is easy both at the ICU bedside and during fieldwork (e.g., in poorly resourced or extreme environment settings using wearable devices). TE-based network measures can be added to ICU digital monitors or portable devices. The current survival prediction and analysis score in the ICU leaves room for foreseeing the potential underlying dysfunctional connections between organ systems in complex diseases. In this case, TE and network indices can be continuously calculated and monitored as a digital value for tracking individuals who require more attention and for making important clinical decisions during patient care. The independence of TEs from SOFA in predicting deterioration and outcomes means that network indices have the potential to be used in conjunction with SOFA and other clinical/laboratory measures in patient care. The independence of TE-based network indices in predicting poor outcomes also provides insight into the pathophysiology of sepsis and emphasizes the importance of an integrated network approach in understanding the mechanisms of dysregulated host responses to infection. Organ system connectivity probably plays an important role in the regulated host physiological response to infection, a concept that is not typically assessed in most cellular/molecular studies, which are carried out using a reductionistic approach (Oyelade et al., 2024).

Thirdly, the findings in Figure 3 optimized the transfer entropy calculation by demonstrating that TE (SpO_2_ → HR), TE (HR → RR) and TE (RR → HR) reaches a plateau at a time lag of approximately 5 seconds and remains stable afterward. This finding is interesting and aligns with previous reports that attempted to estimate the memory length within the cardiorespiratory system (Shirazi et al., 2013). In the context of physiological time-series, memory is a statistical feature that persists for a period and distinguishes the time-series from a random, or memory-less, process (Shirazi et al., 2013). Shirazi et al. developed a method for quantifying memory in physiological time-series and reported that the memory length is estimated to be around 5 to 25 seconds in the cardiorespiratory system in both health and disease (Shirazi et al., 2013). This means any intrinsic perturbation within the physiological system would affect the system for a limited time before the effect dissipates. This limited memory length makes the system more controllable, as prolonged memory can impair the adaptability of the physiological system (Mazloom et al., 2014; Taghipour et al., 2016). Furthermore, a time lag of 5 seconds also represents approximately two respiratory cycles, which aligns with the known physiological interaction between RR and HR within this time frame (e.g., respiratory sinus arrhythmia).

In the analysis of mortality and deterioration prediction, we found that only two directed transfer entropy values showed a consistent pattern of significance for all statistical analyse were HR → RR and RR → HR. In the context of HR → RR and RR → HR, a study of directional coupling between the cardio-respiratory system may explain the clinical significance of transfer entropy. In a recent study, Borovkova et al. revealed the presence of bidirectional couplings between cardiac and respiratory cycles across all age groups in healthy participants (Borovkova et al., 2022). Their findings showed that the coupling from respiration to the parasympathetic control of HR is stronger than the coupling in the opposite direction in health. They also suggested that the directed interaction between RR and HR may be disrupted in complex diseases such as sleep apnoea, leading to an increase in the directional coupling from the main heart rhythm to respiration (Borovkova et al., 2022). This interpretation may also apply to sepsis, where the information transfer is disrupted from RR to HR in patients with poor prognoses due to the loss of directional coupling. Our study indicates that both TE (HR → RR) and TE (RR → HR) are reduced in non-surviving patients with sepsis compared to survivors. However, a full interpretation of these findings awaits further research involving physiological network mapping in health as well as transition from health to disease. We wondered if TE (RR → HR) shows any correlation with the degree of respiratory sinus arrhythmia and thus measured short-term HRV in this cohort using the Poincaré plot, where SD1 is commonly used as a measure of respiratory sinus arrhythmia (Bhogal et al., 2019). We observed that SD1 exhibits a statistically significant correlation with TE (RR → HR) (data not shown). Further studies are required to elucidate the exact interpretation of TE (HR → RR) and its interaction with TE (RR → HR) in health and disease.

### Limitations

There were important limitations to this study. The principal ones were the small cohort size and the use of only three physiological signals. These related issues were due to the relatively low proportion of patients in MIMIC-III with waveform data; the relatively demanding requirement of 30 minutes simultaneous signals with no missing data; and the *a priori* choice to limit inclusion to a Sepsis-3 cohort to reduce the heterogeneity seen in ICU patients. This lack of appropriate data in MIMIC-III may portend issues with TE measurement in the real world: as probes are removed for toileting or other transfers, it may be difficult to obtain unbroken waveform records of sufficient duration for stable estimation of TE and this may limit its potential as a monitor of health. We wondered if shorter time-series can estimate TE calculated from 30-minute time-series and predict poor outcomes (mortality, deterioration) within this patient population. Therefore, we analysed 20-, 10-, 5-, and 1-minute time-series for the calculation of TEs (see Supplementary material 3). Using Bland-Altman analysis, the results showed that different TEs are subject to varying degrees of bias when shorter time-series are used. The most robust TEs were TE (HR → RR) and TE (RR → HR), where 10- and 20-minute time-series could estimate TEs calculated from 30-minute time-series (Supplementary material 3-B2 and B3). Survival analysis also indicated that TE (HR → RR) and TE (RR → HR) calculated from 20-minute time-series could predict mortality and 48-hour deterioration independently of age, SOFA, mechanical ventilation, and comorbidity (Supplementary material 3C). This finding is promising as it shows that shorter time-series can be used for network mapping, which facilitates clinical translation.

It should also be noted that due to early discharge or death, 55% of 48-hour SOFA scores in this study required imputation. While we used a reasonable method imputation of 48-hour SOFA, our findings on prediction of deterioration may be subject to bias and a larger sample size in future studies could provide more solid evidence for the value to TE-based network mapping in prediction of deterioration in sepsis.

Further limitations were the impact of mechanical ventilation and of excessive supplemental oxygen on the measurement of TE. Both of these factors are partially under the control of the clinician, meaning that measured TE may not always directly reflect the patient’s own physiology. In this study, “mechanical ventilation” was defined as both patients undergoing positive pressure ventilation and those using spontaneous breathing modes. Those who were positive pressure ventilated, and in particular paralysed, may have had very low TE values, even if this ventilation was temporary for patients with relatively normal lung function (for example, postoperatively). Supra-normal oxygen saturation levels were also sometimes seen due to excessive supplemental oxygen, both in ventilated and non-ventilated patients. Accidental excessive oxygen administration is common in real world clinical practice (Palmer et al., 2019), but it may have major effect on TE calculation, as it can result in ceiling oxygen saturation (100%) being recorded for every value in the waveform record. These patients then have low or zero TE edge estimates – as target values can be predicted using past information from the target alone. While our results showed that the prognostic value of TEs was independent of mechanical ventilation, future studies can investigate the effect of respiratory support on TEs further.

## Conclusion

This work has confirmed the potential of transfer entropy measurement as a novel digital biomarker in intensive care. Extension of the current methodology to larger datasets is needed to fully understand the interactions of individual TE edges and the impact of patient confounders and mechanical ventilation on its predictive ability.

## Data Availability

All data produced in the present study are available upon reasonable request to the authors

## Acknowledgements

The authors are grateful to UCL Advanced Research Computing Centre (ARC) for collaboration and support.

## Conflict of interest

None

## Authors contribution

Conceived and designed research (MW, WL, ARM), analysed data (CM, MW, QL, EI, CT, P-YC, AC, ARM), interpreted results of experiments (CM, MW, QL, WL, ARM), prepared figures (CM, MW, CT, QL, ARM), drafted manuscript (MW, QL, ARM), edited and revised manuscript (CM, EI, CT, P-YC, WL), approved final version of manuscript (All authors).

## Ethics statement

MIMIC-III is publicly available to researchers under a data use agreement. The data has been deidentified according to HIPAA standards and the project was approved by the Institutional Review Boards of Beth Israel Deaconess Medical Center and MIT (IRB protocol nos. 2001P001699/14 and 0403000206). Individual patient consent was waived as the project did not affect clinical care and all protected health information was deidentified. The authors involved in data extraction completed mandatory online ethics training at MIT and were credentialed (ID 10304625).

## Notes

### Competing Interest Statement

The authors have declared no competing interest.

### Funding Statement

This study did not receive any funding

### Author Declarations

Institutional Review Boards of Beth Israel Deaconess Medical Center and Massachusetts Institute of Technology (IRB protocol nos. 2001P001699/14 and 0403000206).

