## Supplementary material for "Decreased cardio-respiratory information transfer is associated with deterioration and a poor prognosis in critically ill patients with sepsis"

**Supplementary material 1: Multivariate Cox Regression Analysis for Prediction of 30-day Mortality based on Age, SOFA, Elixhauser, Mechanical Ventilation and different Network Indices:**

A: Multivariate Cox Regression for TE(SpO2🡪HR)

|  | **B** | **SE** | **P-value** | **Exp(B)** | **Confidence Interval (95%)** |
| --- | --- | --- | --- | --- | --- |
| Age | 0.04 | 0.013 | 0.003 | 1.040 | 1.013 – 1.068 |
| SOFA | 0.191 | 0.047 | <0.001 | 1.210 | 1.103 – 1.327 |
| Elixhauser | 0.024 | 0.029 | 0.418 | 1.024 | 0.967 – 1.085 |
| Mechanical ventilation | 1.750 | 0.391 | <0.001 | 5.755 | 2.672 – 12.393 |
| TE(SpO_2_🡪HR) | -2.235 | 1.075 | **0.038** | 0.107 | 0.013 – 0.880 |

B: Multivariate Cox Regression for TE(HR🡪RR)

|  | B | SE | P-value | Exp(B) | Confidence Interval (95%) |
| --- | --- | --- | --- | --- | --- |
| Age | 0.044 | 0.014 | 0.002 | 1.045 | 1.017 – 1.075 |
| SOFA | 0.202 | 0.049 | <0.001 | 1.224 | 1.112 – 1.348 |
| Elixhauser | 0.015 | 0.029 | 0.607 | 1.015 | 0.959 – 1.075 |
| Mechanical ventilation | 1.372 | 0.038 | <0.001 | 3.948 | 1.874 – 8.315 |
| TE(HR🡪RR) | -1.925 | 0.889 | **0.030** | 0.146 | 0.026 – 0.832 |

C: Multivariate Cox Regression for TE(RR🡪HR)

|  | B | SE | P-value | Exp(B) | Confidence Interval (95%) |
| --- | --- | --- | --- | --- | --- |
| Age | 0.038 | 0.014 | 0.005 | 1.039 | 1.012 – 1.067 |
| SOFA | 0.192 | 0.048 | <0.001 | 1.212 | 1.104 – 1.330 |
| Elixhauser | 0.017 | 0.029 | 0.547 | 1.018 | 0.961 – 1.077 |
| Mechanical ventilation | 1.787 | 0.393 | <0.001 | 5.969 | 2.761 – 12.903 |
| TE(RR🡪HR) | -2.151 | 1.030 | **0.037** | 0.116 | 0.015 – 0.876 |

E: Multivariate Cox Regression for Indegree of HR

|  | **B** | **SE** | **P-value** | **Exp(B)** | **Confidence Interval (95%)** |
| --- | --- | --- | --- | --- | --- |
| Age | 0.039 | 0.014 | 0.004 | 1.039 | 1.012 – 1.067 |
| SOFA | 0.190 | 0.047 | <0.001 | 1.209 | 1.102 – 1.326 |
| Elixhauser | 0.021 | 0.029 | 0.464 | 1.022 | 0.965 – 1.082 |
| Mechanical ventilation | 1.779 | 0.393 | <0.001 | 5.926 | 2.744 – 12.801 |
| Indegree HR | -1.151 | 0.536 | **0.032** | 0.316 | 0.111 – 0.904 |

F: Multivariate Cox Regression for Outdegree SpO2

|  | **B** | **SE** | **P-value** | **Exp(B)** | **Confidence Interval (95%)** |
| --- | --- | --- | --- | --- | --- |
| Age | 0.044 | 0.014 | 0.002 | 1.045 | 1.017 – 1.075 |
| SOFA | 0.198 | 0.048 | <0.001 | 1.218 | 1.109 – 1.338 |
| Elixhauser | 0.026 | 0.030 | 0.389 | 1.026 | 0.968 – 1.088 |
| Mechanical ventilation | 1.398 | 0.374 | <0.001 | 4.046 | 1.943 – 8.424 |
| Outdegree SpO_2_ | -1.575 | 0.663 | **0.017** | 0.207 | 0.056 – 0.759 |

G: Multivariate Cox Regression for Outdegree HR

|  | **B** | **SE** | **P-value** | **Exp(B)** | **Confidence Interval (95%)** |
| --- | --- | --- | --- | --- | --- |
| Age | 0.044 | 0.014 | 0.002 | 1.046 | 1.018 – 1.075 |
| SOFA | 0.198 | 0.048 | <0.001 | 1.227 | 1.117 – 1.348 |
| Elixhauser | 0.026 | 0.030 | 0.389 | 1.022 | 0.966 – 1.082 |
| Mechanical ventilation | 1.255 | 0.390 | <0.001 | 3.506 | 1.632 – 7.534 |
| Outdegree HR | -1.362 | 0.624 | **0.029** | 0.256 | 0.075 – 0.87 |

H: Multivariate Cox Regression for Outdegree RR

|  | **B** | **SE** | **P-value** | **Exp(B)** | **Confidence Interval (95%)** |
| --- | --- | --- | --- | --- | --- |
| Age | 0.041 | 0.013 | 0.002 | 1.042 | 1.015 – 1.069 |
| SOFA | 0.199 | 0.048 | <0.001 | 1.220 | 1.111 – 1.339 |
| Elixhauser | 0.023 | 0.029 | 0.434 | 1.023 | 0.967 – 1.082 |
| Mechanical ventilation | 1.450 | 0.370 | <0.001 | 4.262 | 2.062 – 8.808 |
| Outdegree RR | -1.462 | 0.698 | **0.036** | 0.232 | 0.059 – 0.911 |

**Supplementary material 2, Multivariate Cox Regression Analysis for Prediction of 48-hour Deterioration based on Age, SOFA, Elixhauser, Mechanical Ventilation and different Network Indices:**

A: Multivariate Cox Regression for TE (HR🡪RR):

|  | **B** | **SE** | **P-value** | **Exp(B)** | **Confidence Interval (95%)** |
| --- | --- | --- | --- | --- | --- |
| Age | 0.044 | 0.014 | 0.002 | 1.045 | 1.017 – 1.075 |
| SOFA | 0.202 | 0.049 | <0.001 | 1.224 | 1.112 – 1.348 |
| Elixhauser | 0.015 | 0.029 | 0.607 | 1.015 | 0.959 – 1.075 |
| Mechanical ventilation | 1.373 | 0.380 | <0.001 | 3.948 | 1.874 – 8.315 |
| TE(HR🡪RR) | -1.925 | 0.889 | **0.030** | 0.146 | 0.026 – 0.832 |

B: Multivariate Cox Regression for TE (RR🡪HR):

|  | **B** | **SE** | **P-value** | **Exp(B)** | **Confidence Interval (95%)** |
| --- | --- | --- | --- | --- | --- |
| Age | 0.038 | 0.014 | 0.005 | 1.039 | 1.012 – 1.067 |
| SOFA | 0.192 | 0.048 | <0.001 | 1.212 | 1.104 – 1.330 |
| Elixhauser | 0.017 | 0.029 | 0.547 | 1.018 | 0.961 – 1.077 |
| Mechanical ventilation | 1.787 | 0.393 | <0.001 | 5.969 | 2.761 – 12.903 |
| TE(RR🡪HR) | -2.151 | 1.030 | **0.037** | 0.116 | 0.015 – 0.876 |

C: Multivariate Cox Regression for Outdegree of SpO2:

|  | **B** | **SE** | **P-value** | **Exp(B)** | **Confidence Interval (95%)** |
| --- | --- | --- | --- | --- | --- |
| Age | 0.020 | 0.013 | 0.111 | 1.020 | 0.995 – 1.046 |
| SOFA | 0.092 | 0.050 | 0.065 | 1.096 | 0.994 – 1.208 |
| Elixhauser | -0.034 | 0.036 | 0.348 | 0.967 | 0.900 – 1.038 |
| Mechanical ventilation | 0.853 | 0.379 | 0.024 | 2.346 | 1.117 – 4.927 |
| Outdegree SpO_2_ | -1.556 | 0.710 | **0.028** | 0.211 | 0.052 – 0.847 |

**Supplementary material 3: The Effect Network Indices Calculated from Shorter Time-series (20, 10, 5, 2, 1 minute) of physiological signals on Prediction of 30-day Mortality and 48-hour Deterioration**

**Method:**

For each patient, transfer entropies were calculated using shorter time series of HR, RR, and SpO2 data. The length of time series used were 20 minutes, 10 minutes, 5 minutes, 2 minutes, and 1 minute.

Bland-Altman plots were used to identify bias in transfer entropy of time series of 20, 10, 5, 2, and 1 minutes compared to the 30-minute transfer entropy values, linear regression analysis was used to test for statistical significance (p<0.05) of the bias for the intercept and slope.

Mono-variate and multivariate (covariates SOFA, Elixhauser score, mechanical ventilation status, and age) Cox regression was used for estimation of hazard ratios with 95% confidence intervals. This was used to test for prediction of mortality and deterioration. ROC curve analysis for mortality was used to find the AUC for transfer entropies of different time-series. These analyses were performed on TE (HR🡪RR), TE (RR🡪HR) and TE (SpO2🡪HR), as they were found to be significant predictors of mortality using time-series length of 30 minutes.

**Results:**

A: The Summary of Bland Altman Plots for Different Transfer Entropies

A.1

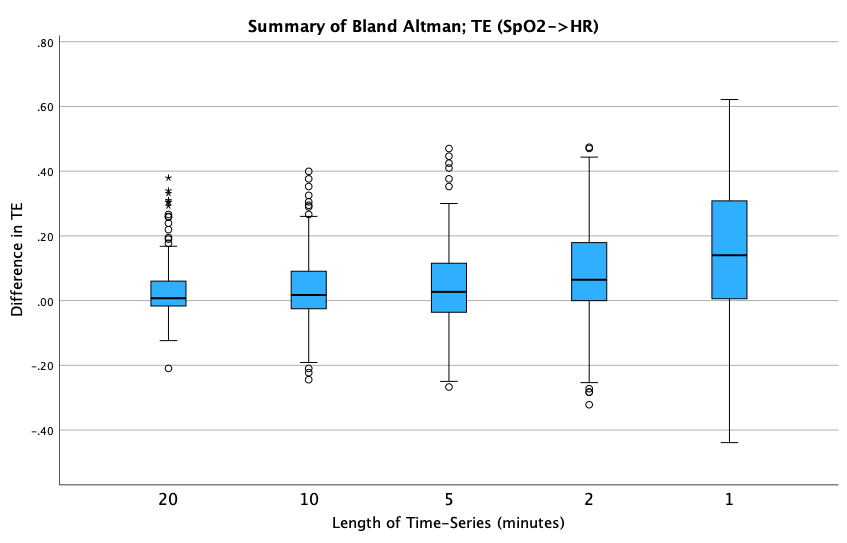

A.2

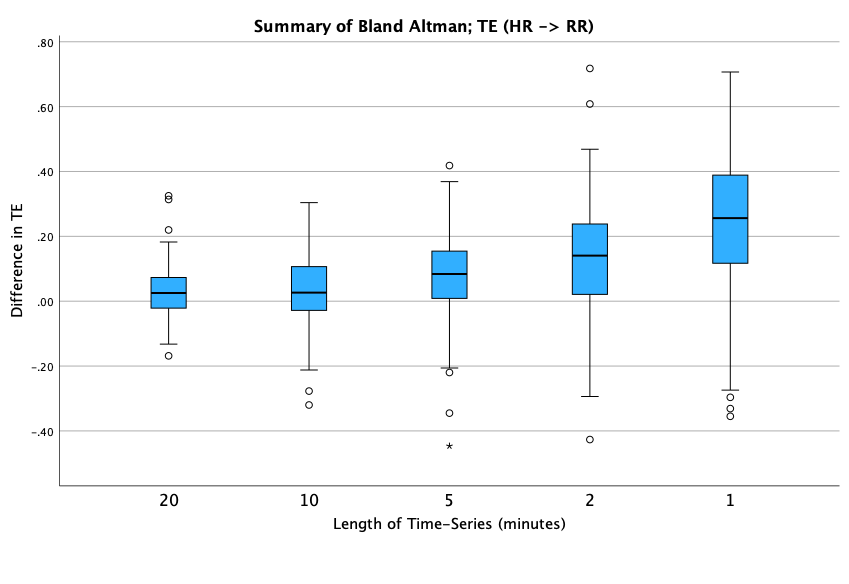

A.3

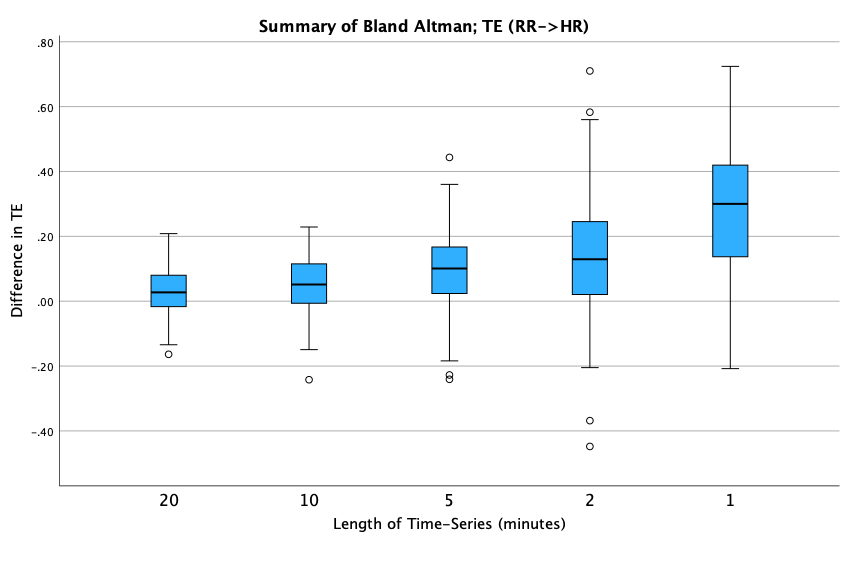

B: The Results of Bland Altman Analysis to Assess Bias using Linear Regression Analysis

B.1: TE (HR 🡪 SpO_2_)

| Time (min) | **Constant B (95% CI)** | **P-value** | **Slope B (95% CI)** | **P-value** |
| --- | --- | --- | --- | --- |
| 20 | 0.022 (-0.006, 0.050) | 0.124 | 0.037 (-0.041, 0.116) | 0.350 |
| 10 | 0.038 (0.005, 0.072) | **0.024** | 0.009 (-0.087, 0.104) | 0.869 |
| 5 | 0.049 (0.008, 0.090) | **0.020** | -0.001 (-0.121, 0.118) | 0.981 |
| 2 | 0.079 (0.034, 0.124) | **<0.001** | 0.017 (-0.120, 0.154) | 0.808 |
| 1 | 0.156 (0.103, 0.209 | **<0.001** | -0.029 (0.209, 0.150) | 0.747 |

B.2: TE (HR 🡪 RR)

| Time (min) | **Constant B (95% CI)** | **P-value** | **Slope B (95% CI)** | **P-value** |
| --- | --- | --- | --- | --- |
| 20 | 0.019 (-0.014, 0.053) | 0.262 | 0.022 (-0.043, 0.087) | 0.501 |
| 10 | 0.007 (-0.040, 0.054) | 0.771 | 0.054 (-0.038, 0.146) | 0.250 |
| 5 | 0.005 (-0.054, 0.064) | 0.886 | 0.160 (0.040, 0.280) | **0.009** |
| 2 | 0.067 (-0.130, 0.147) | 0.102 | 0.148 (-0.027, 0.322) | 0.096 |
| 1 | 0.281 (0.190, 0.373) | **<0.001** | -0.091 (-0.317, 0.136) | 0.431 |

B.3: TE (RR 🡪 HR)

| Time (min) | **Constant B (95% CI)** | **P-value** | **Slope B (95% CI)** | **P-value** |
| --- | --- | --- | --- | --- |
| 20 | 0.018 (-0.015, 0.051) | 0.294 | 0.024 (-0.042, 0.091) | 0.470 |
| 10 | 0.028 (-0.010, 0.066) | 0.150 | 0.040 (-0.038, 0.118) | 0.314 |
| 5 | 0.082 (0.029, 0.135) | **0.003** | 0.032 (-0.083, 0.146) | 0.548 |
| 2 | 0.191 (0.118, 0.264) | **<0.001** | -0.132 (-0.296, 0.033) | 0.116 |
| 1 | 0.329 (0.251, 0.407) | **<0.001** | -0.151 (-0.358, 0.056) | 0.153 |

B.4: TE (SpO_2_ 🡪 RR)

| Time (min) | **Constant B (95% CI)** | **P-value** | **Slope B (95% CI)** | **P-value** |
| --- | --- | --- | --- | --- |
| 20 | 0.047 (0.007, 0.087) | **0.021** | -0.032 (-0.102, 0.037) | 0.357 |
| 10 | 0.027 (-0.024, 0.078) | 0.301 | 0.023 (-0.066, 0.112) | 0.616 |
| 5 | 0.037 (-0.033, 0.107) | 0.296 | 0.065 (-0.061, 0.191) | 0.312 |
| 2 | 0.121 (0.025, 0.218) | **0.014** | 0.033 (-0.152, 0.219) | 0.726 |
| 1 | 0.337 (0.233, 0.442) | **<0.001** | -0.158 (-0.388, 0.072) | 0.177 |

B.5: TE (SpO_2_ 🡪 HR)

| Time (min) | **Constant B (95% CI)** | **P-value** | **Slope B (95% CI)** | **P-value** |
| --- | --- | --- | --- | --- |
| 20 | 0.034 (-0.006, 0.074) | 0.096 | 0.016 (-0.061, 0.093) | 0.684 |
| 10 | 0.065 (0.020, 0.110) | **0.005** | -0.010 (-0.099, 0.078) | 0.817 |
| 5 | 0.116 (0.058, 0.175) | **<0.001** | -0.028 (-0.149, 0.093) | 0.650 |
| 2 | 0.232 (0.152, 0.311) | **<0.001** | -0.187 (-0.357, -0.016) | **0.032** |
| 1 | 0.409 (0.316, 0.502) | **<0.001** | -0.398 (-0.623, -0.173) | **0.001** |

B.6: TE (RR 🡪 SpO2)

| Time (min) | **Constant B (95% CI)** | **P-value** | **Slope B (95% CI)** | **P-value** |
| --- | --- | --- | --- | --- |
| 20 | 0.033 (0.004, 0.062) | **0.028** | 0.022 (-0.055, 0.099) | 0.580 |
| 10 | 0.040 (0.004, 0.075) | **0.030** | 0.027 ( -0.069, 0.122) | 0.581 |
| 5 | 0.047 (0.002, 0.093) | **0.042** | 0.046 (-0.078 ,0.171) | 0.465 |
| 2 | 0.102 (0.051, 0.153) | **<0.001** | -0.018 (-0.164, 0.127) | 0.803 |
| 1 | 0.148 (0.090, 0.205) | **<0.001** | 0.101 (-0.085, 0.287) | 0.286 |

C: Comparison of Different Time-series Lengths on Prediction of 30-day Mortality and 48-hour Deterioration. Highlighted boxes indicate statistically significant results.

**C.1: TE (HR 🡪 RR)**

|  | **Prediction of Mortality** | | | **Prediction of Deterioration** | |
| --- | --- | --- | --- | --- | --- |
| Time (min) | **Hazard Ratio (95%CI)** | **Multivariate Hazard Ratio (95% CI)** | **ROC (AUC 95% CI)** | **Hazard Ratio (95% CI)** | **Multivariate Hazard Ratio (95% CI)** |
| 30 | 0.035 (0.007 – 0.166) ***** | 0.146 (0.026–0.832) ***** | 0.724 (0.622 – 0.827) ***** | 0.058 (0.011 – 0.304) ***** | 0.156 (0.026 – 0.940) ***** |
| 20 | 0.030 (0.006 – 0.159) ***** | 0.120 (0.019 – 0.759) ***** | 0.707 (0.603 – 0.811) ***** | 0.048 (0.008 – 0.271) ***** | 0.032 ( 0.018 – 0.842) ***** |
| 10 | 0.036 (0.006-0.205) ***** | 0.120 (0.021 – 0.680 ) ***** | 0.698 (0.591 – 0.804) ***** | 0.083 (0.013 – 0.517) ***** | 0.178 (0.028 – 1.127) |
| 5 | 0.039 (0.006 - 0.270) ***** | 0.120 (0.015 – 0.948) ***** | 0.678 (0.571 – 0.785) ***** | 0.149 (0.018 – 1.232) | 0.366 (0.042 – 3.156) |
| 2 | 0.132 (0.015 – 1.194) | 0.529 (0.055 – 5.045) | 0.600 (0.482 – 0.718) | 0.081 (0.008 – 0.766) | 0.238 (0.025 – 2.290) |
| 1 | 0.379 (0.059 – 2.450) | 0.506 (0.076 – 3.372) | 0.562 (0.449 – 0.675) | 0.490 (0.072 – 3.331) | 0.412 (0.060 – 2.845) |

**C.2: TE ( RR 🡪 HR)**

|  | **Prediction of Mortality** | | | **Prediction of Deterioration** | |
| --- | --- | --- | --- | --- | --- |
| Time (min) | **Hazard Ratio (95%CI)** | **Multivariate Hazard Ratio (95% CI)** | **ROC (AUC 95% CI)** | **Hazard Ratio (95% CI)** | **Multivariate Hazard Ratio (95% CI)** |
| 30 | 0.068 (0.012 – 0.393) ***** | 0.116 (0.015 – 0.876) ***** | 0.635 (0.518 – 0.715) ***** | 0.131 (0.020 – 0.843) ***** | 0.163 (0.020 – 1.353) |
| 20 | 0.029 (0.005 – 0.176) ***** | 0.071 (0.009 – 0.527) ***** | 0.667 (0.551 – 0.783) ***** | 0.070 (0.010 – 0.474) ***** | 0.104 (0.012 – 0.875) ***** |
| 10 | 0.039 (0.007 – 0.238) ***** | 0.073 (0.009 – 0.560) ***** | 0.650 (0.532 – 0.768) ***** | 0.104 (0.015 – 0.720) ***** | 0.124 (0.014 – 1.123) |
| 5 | 0.034 (0.005 – 0.221) ***** | 0.035 (0.005 – 0.248) ***** | 0.689 (0.587 – 0.792) ***** | 0.129 (0.017 – 0.986) ***** | 0.153 (0.018 – 1.325) |
| 2 | 0.194 ( 0.033 – 1.124) ***** | 0.174 (0.027 – 1.128) | 0.597 (0.484 – 0.709) | 0.189 (0.030 –1.186) | 0.238 (0.034 – 1.662) |
| 1 | 0.093 (0.011 – 0.780) ***** | 0.130 (0.016 – 1.050) | 0.626 (0.521 – 0.732) ***** | 0.254 (0.031 – 2.061) | 0.405 (0.052 – 3.151) |

**C.3: TE ( SpO_2_ 🡪 HR)**

|  | **Prediction of Mortality** | | | **Prediction of Deterioration** | |
| --- | --- | --- | --- | --- | --- |
| Time (min) | **Hazard Ratio (95%CI)** | **Multivariate Hazard Ratio (95% CI)** | **ROC (AUC 95% CI)** | **Hazard Ratio (95% CI)** | **Multivariate Hazard Ratio (95% CI)** |
| 30 | 0.072 (0.012 – 0.421) ***** | 0.107 (0.013 – 0.880) ***** | 0.658 (0.554 – 0.761) ***** | 0.104 (0.016 – 0.674) ***** | 0.128 (0.014 – 1.178) |
| 20 | 0.046 ( 0.007 – 0.304) ***** | 0.126 (0.015 – 1.095) ***** | 0.659 (0.545 – 0.773) ***** | 0.154 (0.019 – 1.216) ***** | 0.307 (0.031 – 3.009) |
| 10 | 0.042 (0.006 – 0.275) ***** | 0.109 (0.013 – 0.931) ***** | 0.650 (0.537 – 0.764) ***** | 0.155 (0.021 – 1.153) ***** | 0.231 (0.024 – 2.232) |
| 5 | 0.019 (0.003 – 0.113) ***** | 0.011 (0.001 – 0.101) ***** | 0.713 (0.613 – 0.814) ***** | 0.085 (0.013 – 0.563) ***** | 0.086 (0.010 - 0.769) ***** |
| 2 | 0.104 (0.018 – 0.585) ***** | 0.072 (0.010 – 0.537) ***** | 0.605 (0.492 – 0.718) ***** | 0.360 (0.058 – 2.239) | 0.483 (0.066 – 3.541) |
| 1 | 0.934 (0.180 – 4.844) | 0.833 ( 0.144 – 4.837) | 0.500 (0.383 -0.617) | 1.407 (0.255 – 7.769) | 1.585 (0.256 – 9.817) |
